# The ground level ozone concentration is inversely correlated with the number of COVID-19 cases in Warsaw, Poland

**DOI:** 10.1101/2020.09.20.20198366

**Authors:** Oskar Wiśniewski, Wiesław Kozak, Maciej Wiśniewski

## Abstract

COVID-19, which is a consequence of infection with the novel viral agent SARS-CoV-2, first identified in China (Hubei Province), has been declared a pandemic by the WHO. As of September 10, 2020, over 70,000 cases and over 2,000 deaths have been recorded in Poland. Of the many factors contributing to the level of transmission of the virus, the weather appears to be significant. In this work we analyse the impact of weather factors such as temperature, relative humidity, wind speed and ground level ozone concentration on the number of COVID-19 cases in Warsaw, Poland. The obtained results show an inverse correlation between ground level ozone concentration and the daily number of COVID-19 cases.

## 1. INTRODUCTION

COVID-19 is a disease caused by the novel viral agent SARS-CoV-2. It was first identified and described in central China (Wuhan, Hubei Province) in December 2019. The World Health Organization announced COVID-19 outbreak a pandemic on March 12, 2020. Until now (September 10, 2020), over 27 million cases and nearly 900,000 deaths caused by this disease were reported worldwide. In Poland, these rates amounted to over 70,000 cases and over 2,000 deaths. Each COVID-19 case follows an individual course. Most infected people develop symptoms that can be described as mild to moderate. Such people recover without hospitalization. However, severe symptoms of the disease are: shortness of breath, high fever, cough, loss of sense of smell, and muscle pain. The mean incubation period of SARS-CoV-2 was estimated to be 6.4 days, ranging from 2.1 to 11.1 days (Lauer et al. 2020).

The first case of the novel coronavirus in Poland was reported on March 4, 2020 (in the western region of the country). The Polish government declared the state of epidemic threat on March 14, 2020 and the state of epidemic was declared on March 20, 2020. At this time, many restrictions were introduced, such as the obligation to wear face masks, a ban on gathering and moving in groups, and the closure of certain service points. It was also recommended not to travel for Easter and the long weekend (in the beginning of May).

It is widely known that weather conditions can influence the transmission of various viruses. Bi et al. (2007) found an inverse association between the number of daily cases of SARS and ambient temperature, and a positive association with air pressure during the epidemic of 2003 in Beijing and Hong Kong. Moreover, several epidemiological analyses identified absolute humidity and temperature as climatic predictors of influenza virus transmission in temperate regions of the world (Lowen and Steel, 2014). The mechanisms by which weather factors shaping the viral transmissibility remain unclear. Possible explanations consider the instability of some viruses at high temperatures, better adhesion to objects at higher humidity, and higher transmission during strong winds.

In the present report, we analyse the impact of daily average temperature, relative humidity, wind speed and ground level ozone concentration on the number of daily COVID-19 cases. As far as we know, our work is the first to consider the impact of weather factors on SARS- CoV-2 transmission in Poland.

## 2. METHODS

### 2.1. Study area

Warsaw is the capital of the Republic of Poland. It is located in the central-eastern part of the country (52° 14’ 13.3764’’ N, 21° 1’ 3.1152’’ E), in the area of humid continental climate (Köppen climate classification). According to data from December 31, 2019, it is inhabited by 1,790,658 people and its area covers 517.24 km^2^ (including the Vistula river).

### 2.2. Data collection

Data on the daily number of cases (confirmed by the RT-PCR method) between April 7, 2020 and June 7, 2020 were obtained from the official reports of the District Sanitary- Epidemiological Station in Warsaw. The weather data from April 1, 2020 to June 1, 2020: daily average temperature [°C], relative humidity [%] and wind speed [m/s] for the period of two months were obtained from the publicly available repository of Polish Institute of Meteorology and Water Management (*Warszawa-Bielany* weather monitoring station). The daily ground level ozone (O3) [µg/m^3^] measurements were obtained from the publicly available database of the Chief Inspectorate of Environmental Protection (*Warszawa- Chroscickiego* air pollution monitoring station). Data on confirmed daily cases were paired with weather data seven days earlier (due to reasons described below).

### 2.3. Data analysis

Since the data were not normally distributed, Spearman rank correlation test was used to examine the relationship between particular weather factors and daily COVID-19 cases.

## 3. RESULTS AND DISCUSSION

Between April 7, 2020 and June 7, 2020 a total number of 917 COVID-19 cases were reported in Warsaw. The lowest daily average temperature recorded in the period between April 1, 2020 and June 1, 2020 was 3.9 °C, while the highest daily average temperature was 20.1 °C. The lowest daily relative humidity recorded was 35.5%, while the highest daily relative humidity was 87%. The lowest wind speed recorded was 1 m/s, while the highest wind speed was 5.7 m/s. The lowest daily average ground level O3 concentration recorded was 53.9 µg/m3, while the highest ground level O3 concentration was 97.9 µg/m3. Changes of respective values over time are presented in Figure 1.

**Figure 1.**
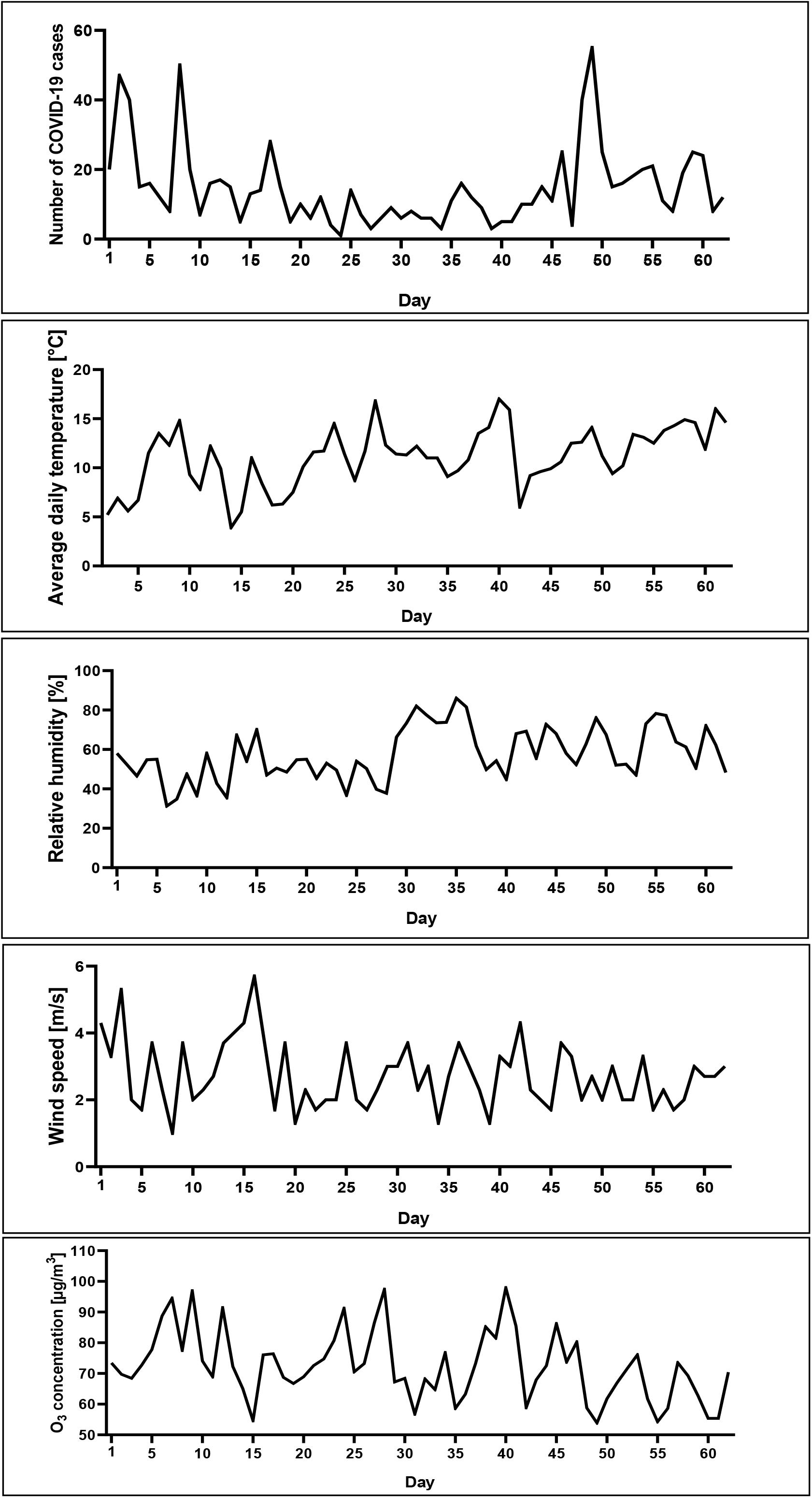
Daily cases of the COVID-19, daily average temperature [°C], relative humidity [%], wind speed [m/s] and average ground level ozone concentration [µg/m^3^] in Warsaw, Poland from April 7 to June 7, 2020 (number of COVID-19 cases) and April 1 to June 1, 2020 (weather variables).

The obtained data indicate that only the average daily ambient ozone concentration is inversely correlated with the number of COVID-19 cases (Tab. 1). Pairing of the daily case numbers with the weather conditions 7 days earlier is in our opinion corresponds more to reality than comparing the case numbers and the weather conditions on the same day. It was done due to the average incubation period of the coronavirus and the average 24-hour waiting period for the RT-PCR test result in Poland.

**Table 1.** Spearman correlation coefficients (*ρ*) and their P-values between COVID-19 cases and weather variables.

| Weather variable | Spearman's $\rho$ | P-value |
| --- | --- | --- |
| Average daily temperature [°C] | -0.1195 | 0.3589 |
| Relative humidity [%] | 0.0389 | 0.7636 |
| Wind speed [m/s] | 0.1355 | 0.2937 |
| Ground level ozone concentration [$\mu\text{g}/\text{m}^3$] | -0.2997* | 0.0180 |

\* Correlation is significant at the 0.05 level (two-tailed).

There is a number of publications describing the impact of weather conditions on the spread of the novel coronavirus. Tosepu et al. (2020) found that only daily average temperature was significantly correlated with number of COVID-19 cases in Jakarta. Luo et al. (2020) reported a positive correlation between absolute humidity and case increase, and a weak negative correlation between weather temperature and case increase in several regions in Asia. A study on the effects of temperature and humidity on the daily new cases and new deaths of COVID- 19 in 166 countries revealed that both factors were negatively related to the daily new cases and daily new deaths of COVID-19 (Wu et al., 2020). Unfortunately, there are no detailed local data on this subject from Central and Eastern Europe.

Only a few papers describe the effects of ambient ozone concentration on COVID-19. A recent paper from China shows that the COVID-19 transmissibility could be negatively associated with ambient ozone (Ran et al., 2020). On the contrary, a positive correlation was found between COVID-19 infections and ground level ozone in Milan, Italy (Zoran et al., 2020). An analysis performed by Adhikari and Yin (2020) showed that daily average temperature, daily maximum eight-hour ozone concentration, average relative humidity, and cloud percentages were significantly and positively associated with new confirmed cases related to COVID-19, but none of these variables showed significant associations with new deaths related to COVID-19. It should be noted that from a biological point of view, ozone is generally considered as an antiviral agent that damages the viral capsid and upsets its reproductive cycle (Elvis and Ekta, 2011).

It must be emphasized that our research has some limitations. Warsaw, as the largest city in the country, is the main communication hub, and the mobility of its inhabitants is at a high level - many of them are commuters from surrounding regions. In addition, the inhabitants are characterized by different hygienic habits, lifestyle, profession, etc. Due to this fact, social factors, in addition to weather conditions, play a significant role in the spread of the virus.

## 4. CONCLUSIONS

Of the factors studied, only ozone concentration seems to have an impact on the number of new COVID-19 cases. Further research is needed, for example in other Polish cities, to characterize the remaining weather factors that may influence the spread of the virus.

## Data Availability

All data are publicly available

## ACKNOWLEDGEMENTS

The authors are grateful to Dr. Maciej Wachulec, who supported and encouraged this research.

